# Multicenter Comparison of AI Deep Learning Reconstruction, Iterative Reconstruction, and Filtered Back Projection for Coronary Artery Calcification Scoring

**DOI:** 10.1101/2024.10.30.24316447

**Authors:** Alena R. Winkler, June D. Campos, Mark L. Winkler

**Affiliations:** Department of Radiology, Steinberg Diagnostic Medical Imaging, Las Vegas, Nevada, United States of America

**Author notes:** Department of Radiology, Steinberg Diagnostic Medical Imaging, 7301 Peak Dr., Ste 200, Las Vegas, NV 89128. These authors contributed equally to this work.

## Abstract

**Objective:** To validate the feasibility of AI Deep Learning Reconstruction for Coronary Artery Calcification Scoring in order to decrease radiation exposure on a 4cm detector CT scanner. This is the first such validation on devices that are most commonly utilized for this procedure.

**Methods:** Data from 105 consecutive patients referred for Coronary Artery Calcification Scoring (CACS) in 4 centers was reconstructed with Filtered Back Projection (FBP), Iterative Reconstruction (Hybrid-IR), and AI Deep Learning Reconstruction (AI DLR), and analyzed both quantitatively and qualitatively to determine if AI DLR can be routinely used for this purpose. Additional phantom testing was performed to determine if further dose reduction can be accomplished with AI DLR while maintaining or improving image quality compared to current Hybrid-IR reconstruction.

**Results:** Quantitively, there was excellent agreement between the three reconstructions (FBP, Hybrid IR and AI DLR) with an interclass coefficient of 0.99. The mean CACS for Filtered Back Projection Reconstructions was 111.05. The mean CACS for Hybrid-IR was 91.30. The mean CACS for AI Deep Learning Reconstructions was 93.50. Qualitatively, image quality was consistently better with AI DLR than with Hybrid-IR at both soft tissue and lung windowing. Based on our phantom experiments, AI DLR allows for dose reduction of at least a 37% without any image quality penalty compared to Hybrid-IR.

**Conclusions:** The use of AI DLR for use in CACS on 4 cm coverage CT scanner has been quantitatively and qualitatively validated for use for the first time. AI DLR produces qualitatively and quantitively better image quality than Hybrid-IR at the same dose level, and produces good agreement in categorization of Agatston scores. In vivo and in vitro evaluations show that AI DLR will allow for an at least a 37% further dose reduction on a 4 cm coverage CT scanner.

## INTRODUCTION

Coronary Artery Calcium Scoring (CACS) is a widely available, low cost, non-invasive imaging test which measures the amount of calcified plaque in the coronary arteries. This score is utilized to assess the risk of coronary artery disease, guide lifestyle modification and therapeutic treatment, and monitor disease progression and response to treatment [1,2,3].

A patient’s level of CACS may be described by the Agaston score based on a low-dose CT scan of the heart. The Agaston score quantifies the amount of calcified plaque in the coronary arteries. Each calcified plaque is given a score calculated by multiplying the area of the calcified plaque by a scale factor determined by maximum density of the plaque [4,5]. The patient score is a sum of all individual plaque scores.

A score of 0 indicates no identifiable calcified plaque and indicates a very low (less than 5%) risk of significant obstructive coronary artery disease (CAD). A score of 1-10 indicates minimal calcified plaque and indicates a low (less than 10%) risk of significant obstructive CAD. A score of 11-100 indicates mild calcified plaque and indicates a low to moderate risk of significant obstructive CAD with mild stenosis likely present. A score of 101-400 indicates moderate calcified plaque and indicates a moderate to high risk of significant obstructive CAD with non-obstructive and obstructive disease likely present. A score of over 400 indicates severe diffuse calcified plaque and indicates a high risk of CAD with at least one significant obstruction (greater than 90%) likely present [6].

CACS may be associated with a significant radiation dose, ranging from 0.8 to 10.5mSv [7]. Such radiation doses have been associated with the risks of subsequent tumors [8,9,10,11] and therefore minimization of dose is of paramount importance.

The CACS CT test may be reconstructed with a variety of methods. Hybrid-IR has been proven to provide lower doses [12, 13, 14, 15, 16, 17] and higher image quality [18, 19] than earlier FBP methods. A previous study with a phantom and human subjects scanned with a wide area detector scanner (16 cm) found that DLR significantly reduced image noise but produced no significant differences in measured calcium volumes [20]. In this study quantification of coronary artery calcium was equivalent between FBP, Hybrid-IR and AI-DLR, with AI-DLR having the lowest bias in measured calcium volumes [20].

This study attempts to determine if DLR can supplant Hybrid-IR to then allow for further dose reduction and improved safety, as suggested in prior studies [18,20, 21] on non-wide area detector CT scanner.

## MATERIALS and METHODS

This retrospective study was approved by SDMI’s institutional Ethics Committee with a waiver of informed consent. 105 consecutive patients referred for Coronary Artery Calcium Scoring (CACS) in 4 outpatient centers between March 4, 2024, and June 18, 2024, were studied as part of our institution’s routine quality improvement process. Deidentified records were accessed between March 17 and July 1, 2024, for the purposes of this study.

There were 57 males and 48 females. CACS studies were performed on CT scanners with 80 row, 4 cm detectors (Prime SP, Canon Medical Systems [93 patients] and Serve SP, Canon Medical Systems [12 patients]). Studies were cardiac gated with a step and shoot technique at a rotation time of 0.35s. Tube voltage was 120 kV for all studies. Data was acquired at 0.5mm slice thickness and reconstruction at a 3mm slice thickness.

Automated exposure control was set to a standard deviation of the noise target of 45 (SD=45) based on the Hybrid-IR model at a 3mm reconstructed slice thickness. This standard deviation level was selected from our prior clinical testing as the level at which non-obese (BMI under 30 kg/m^2^) patients would be exposed to a dose under 1 mSv for their CACS exam.

Scanning was performed from 1.5cm above the coronary arteries to 1.5 cm below the left ventricle. Determination of the scan range was assisted by an optical patient positioning system and a low dose 3D localizing scan.

Data was reconstructed utilizing three methods: Filtered Back Projection, Hybrid-IR (AIDR, Standard Level), and AI Deep Learning Reconstruction (AiCE, Standard Level).

Dose was calculated as the product of Dose Length Product multiplied by the Chest k factor of 0.014 as specified by American Association of Physicists in Medicine report of January 2008 [22].

Quantitative and qualitative CACS was performed by two experienced cardiac imagers with a combined total of 41 years of cardiac imaging experience. A Vitrea workstation (Vitrea Advanced Visualization, Canon Medical Systems) was used to generate the Agaston scores.

A qualitative image quality assessment of reconstruction algorithms was done using a soft tissue (L40, W350) and lung (L-650, W1600) window settings. The relative change in image quality was done using a +/- 3 scale, with 0 being no clinical diagnostic difference, +/- 1 being mild clinical diagnostic difference, +/- 2 being moderate clinical diagnostic difference, and +/-3 being significant clinical diagnostic difference with the Hybrid-IR image quality set as the reference level of zero.

Phantom scans to evaluate the effects of the reconstruction algorithm on radiation dose and image noise were performed using a Catphan 500 phantom (Phantom Laboratory, Salem, NY, USA). To evaluate the dose reduction that could be achieved by changing the reconstruction method for a fixed noise target, scans were performed with noise targets ranging from SD = 10 to SD = 60. The phantom was scanned and scan doses recorded with Hybrid-IR set as the reconstruction method and repeated with AI DLR set as the reconstruction method.

To quantitively evaluate the noise levels produced by each reconstruction method, images from all scans were reconstructed with FBP, Hybrid-IR and AI DLR. Image noise was measured at the central three slices of the uniformity section of the phantom, using a circular ROI approximately 100 cm^2^ in size at the center of the image. These measurements provide a quantitative assessment of noise of each reconstruction method at a fixed scan dose, as well as indicators of potential dose reductions at fixed levels of image noise.

Since the data did not follow a normal gaussian distribution the interclass coefficient measure was used to evaluate the agreement between the three different reconstructions. The Fleiss Kappa statistic (κ) was calculated to evaluate the agreement of AI-DLR and FBP with the standard of care reconstruction (Hybrid-IR) [23].

## RESULTS

### QUANTITATIVE

Patient ages ranged from 27 to 77 with a mean age of 55.7 years. Body Mass Indices (BMI) ranged from 16.7 to 39.5 with a mean of 27.5 kg/m^2^.

The mean CACS for Filtered Back Projection Reconstructions was 111.05. The mean CACS for Hybrid-IR was 91.30. The mean CACS for AI Deep Learning Reconstructions was 93.50 (Figure 1).

**Figure 1.**
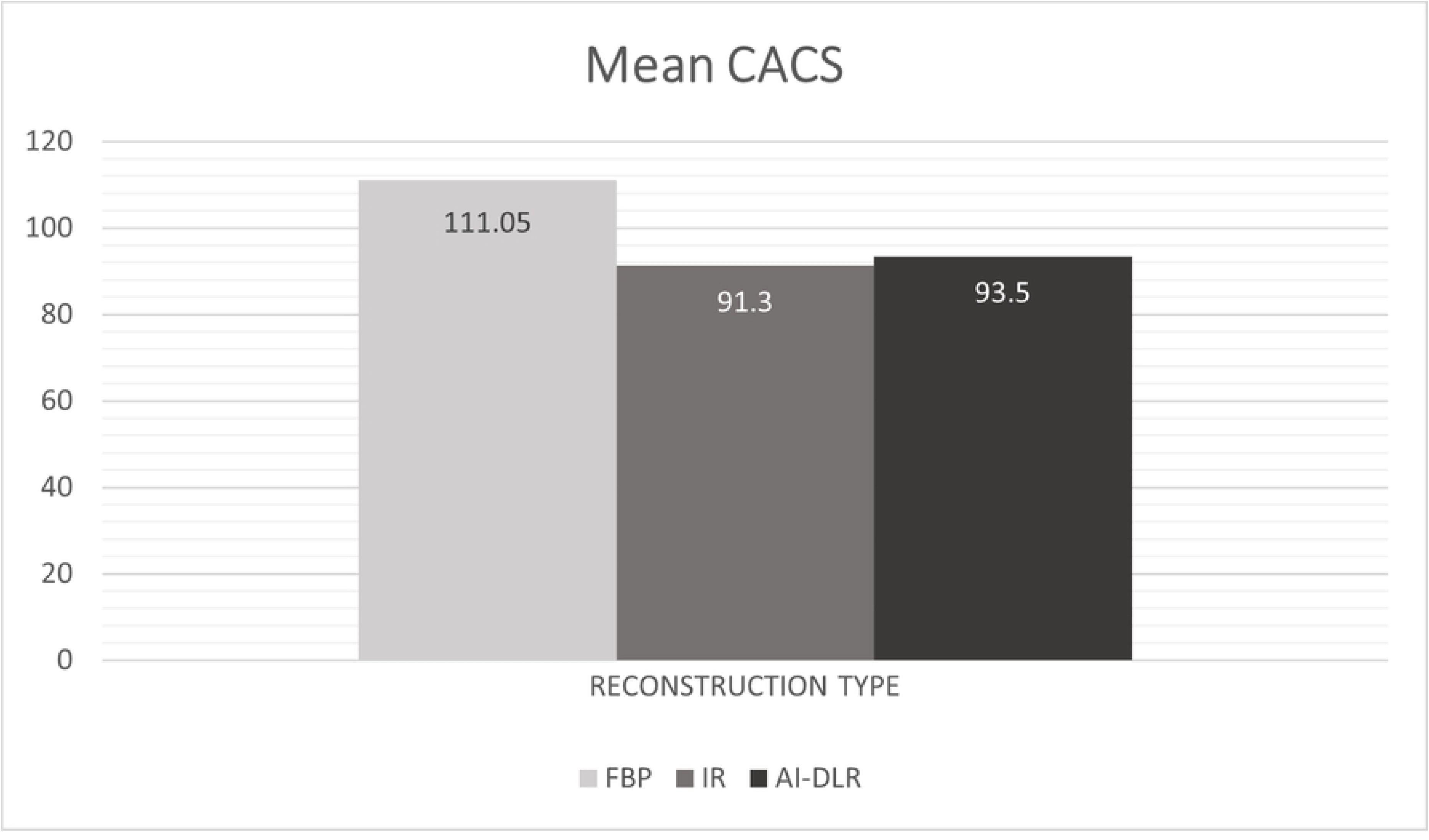
Comparison of Mean CACS across reconstruction types.

All reconstructions FBP, Hybrid-IR and AI -DLR were tested for normality and had D values of 0.3573, 0.3681 and 0.3610 and P <0.0001 which rejects normality of this data. Therefore, an interclass correlation coefficient, which describes the degrees of consistency among measurements, was calculated and found to be 0.99 for the average measures for all 3 reconstructions. This indicates excellent agreement between measurements obtained using the 3 reconstruction methods.

Figure 2 shows the distribution of CACS in each of the risk categories. No coronary calcification (score 0) was seen for 51 patients with FBP, 61 patients with Hybrid-IR, and 58 patients with AI DLR (Figure 2). The kappa statistic (κ) between those patients who were in the CACS category of 0 for AI DLR and Hybrid-IR was 0.94 ± 0.03 while the κ between Hybrid IR and FBP was 0.81 ± 0.05. The two reconstructions (AI DLR and FBP) show strong agreement with Hybrid for those patients with zero calcium.

**Figure 2.**
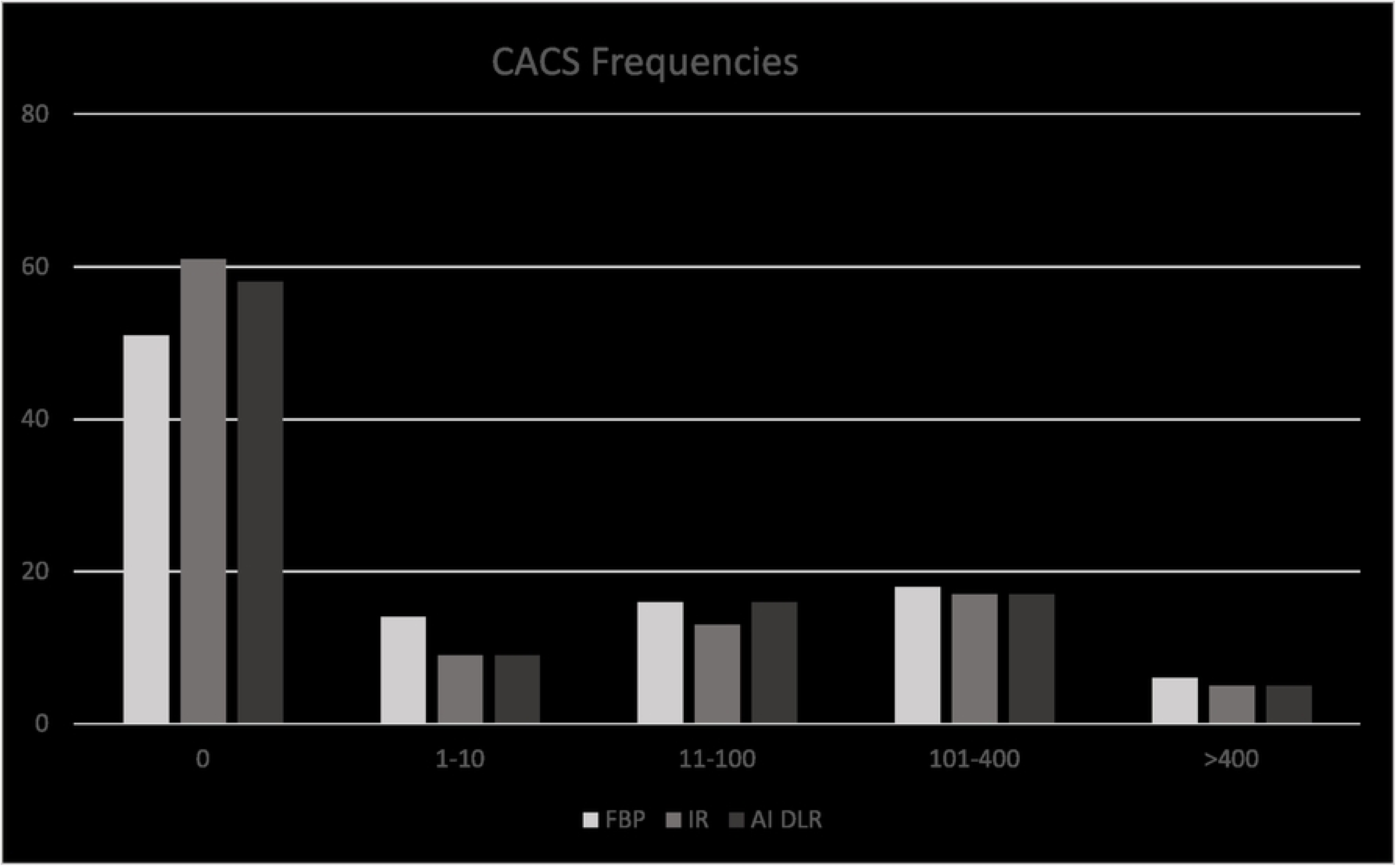
Relative occurrence of CACS values as a function of reconstruction type.

Minimal coronary calcification (score 1-10) was seen for 14 patients with FBP, 9 patients with Hybrid-IR, and 9 patients with AI DLR. The kappa statistic (κ) for AI DL and Hybrid IR was 0.64 ± 0.14 while, the κ between Hybrid IR and FBP was only 0.27 ± 0.14.

Mild coronary calcification (score 11-100) was seen for 16 patients with FBP, 13 patients with Hybrid-IR, and 16 patients with AI DLR. The kappa statistic (κ) for those patients in the mild coronary calcification category between AI DLR and Hybrid-IR was 0.80 ± 0.09, which is a strong agreement while κ between Hybrid IR and FBP was 0.72 ± 0.1.

Moderate coronary calcification (score 101-400) was seen for 18 patients with FBP, 17 patients with Hybrid-IR, and 17 patients with AI DLR. The kappa statistic (κ) for those patients in the moderate coronary calcification category between AI DLR and Hybrid-IR was 0.92 ± 0.05, which is still strong agreement while κ between Hybrid IR and FBP was 0.90 ± 0.06.

Severe coronary calcification (score over 400) was seen for 6 patients with FBP, 5 patients with Hybrid-IR, and 5 patients with AI DLR. The kappa statistic (κ) between AI DLR and Hybrid-IR was 1.0 ± 0.0, which is strong agreement while the κ between Hybrid IR and FBP was 0.9 ± 0.09.

Patient dose was exponentially related to BMI by linear regression analysis and is demonstrated graphically in Figure 3 with a Pearson correlation coefficient of 0.7282 indicating a moderately strong correlation. The dose range was from 0.225 mSv to 3.10 mSv with a mean dose of 0.795 mSv.

**Figure 3.**
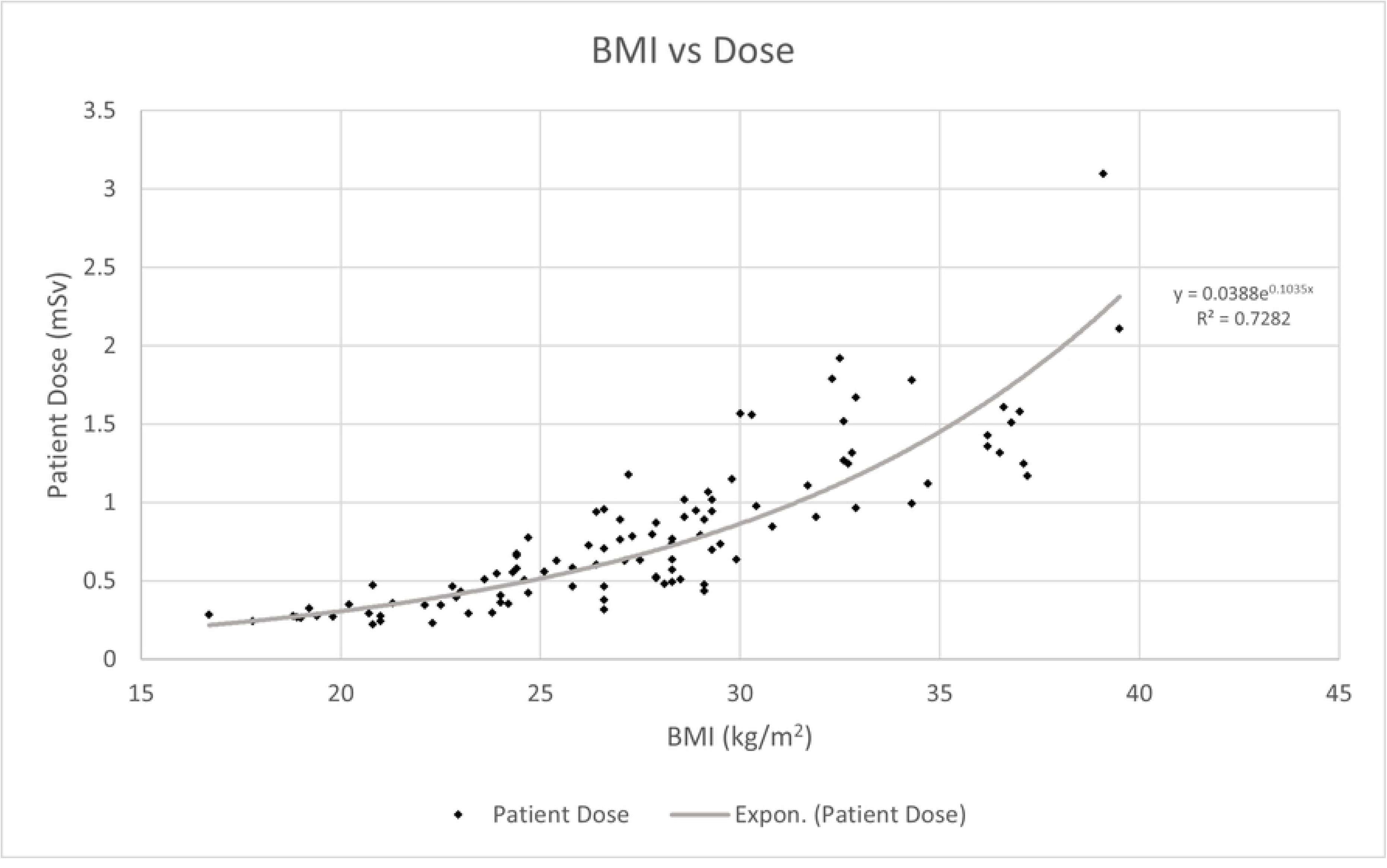
Distribution of patient dose as a function of patient size as represented by patient BMI.

Phantom measurements showed a reduction in CTDI of approximately 30% when the target reconstruction method was changed from Hybrid-IR to AI DLR, but with a fixed noise target (SD). These results are depicted in Figure 4 and Table 1. Table 1 tabulates the CTDI for scans of the Catphan phantom as the noise target was increased when the scan protocol was setup with Hybrid-IR and repeated with AI DLR as the reconstruction. From this table the dose for a noise target of 45 (SD=45) which is our clinical protocol, will reduce the dose by 32% simply by switching the reconstruction being utilized to AI DLR.

**Figure 4.**
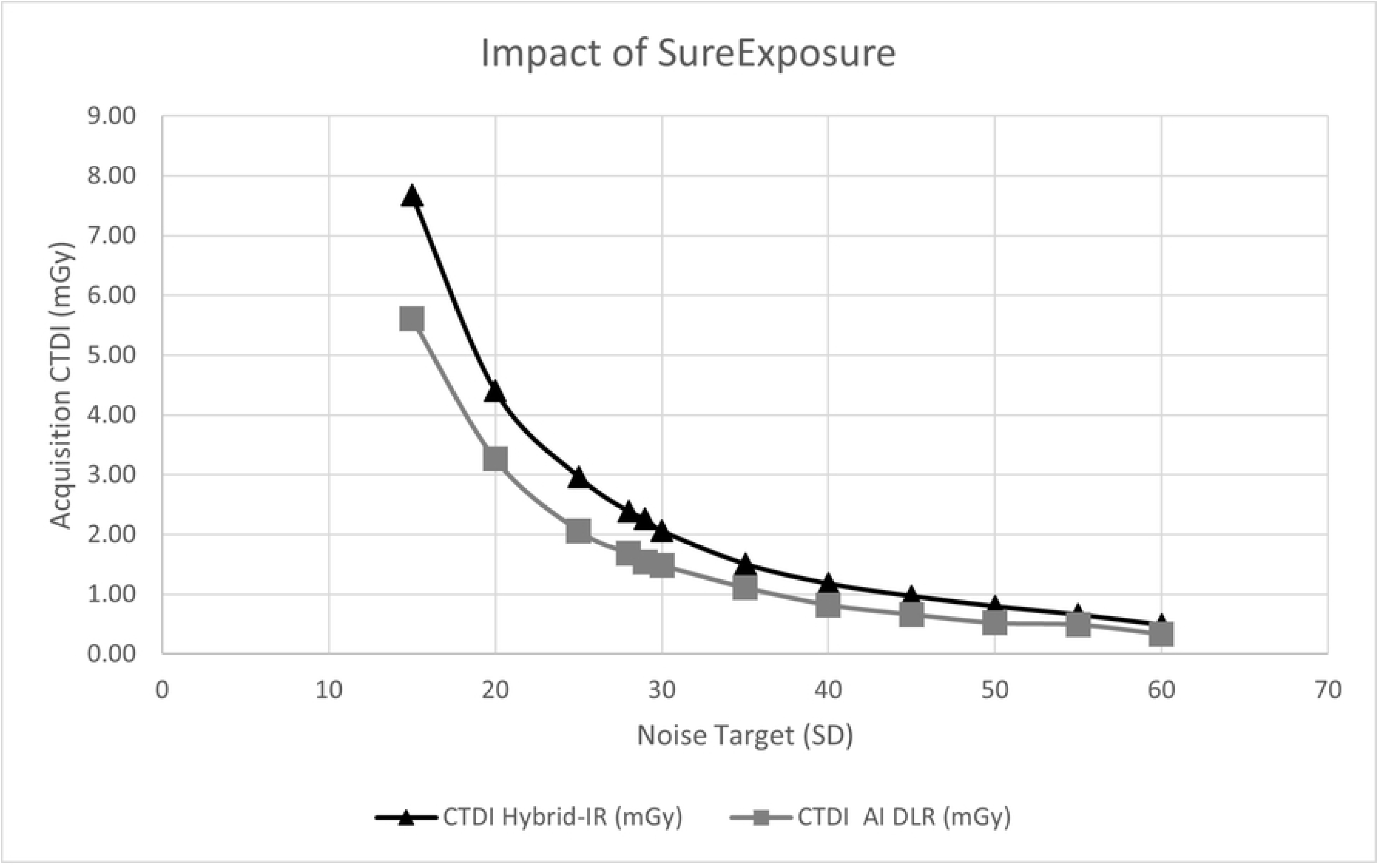
Impact of reconstruction target on dose. For a fixed SD level, switching reconstruction algorithms from Hybrid-IR to AI DLR results in approximately 30% reduction in radiation dose.

**Table 1.** Dose reduction achieved between Hybrid-IR and AI DLR reconstructions.

| Noise Target (SD) | CTDI Hybrid-IR (mGy) | CTDI AI DLR (mGy) | Reduction in Dose |
| --- | --- | --- | --- |
| 15 | 7.68 | 5.61 | 27% |
| 20 | 4.41 | 3.26 | 26% |
| 25 | 2.97 | 2.06 | 31% |
| 28 | 2.39 | 1.69 | 29% |
| 29 | 2.27 | 1.54 | 32% |
| 30 | 2.06 | 1.48 | 28% |
| 35 | 1.51 | 1.11 | 26% |
| 40 | 1.18 | 0.82 | 31% |
| 45 | 0.97 | 0.66 | 32% |
| 50 | 0.80 | 0.52 | 35% |
| 55 | 0.66 | 0.49 | 26% |
| 60 | 0.49 | 0.33 | 33% |

Noise measurements in phantom images were also used to determine the level of dose reduction possible with AI DLR to maintain similar noise levels as Hybrid-IR. Figure 5 shows the effect of the image of reconstruction algorithm on image noise and indicates a dose reduction of at least a 37% would lead no degradation in AI DLR image noise compared to Hybrid IR AI DLR yield lower noise levels compared to FBP or Hybrid-IR. To maintain the noise level attained with Hybrid-IR at a noise target level of 45 (SD=45), a higher noise target (SD = 70) may be used, and this would lead to a dose reduction of approximately 44%. Dashed arrows in Figure 5 illustrates the opportunity for dose reduction. In our phantom experiments, with noise target set at SD = 28, a CTDI of 2.4 mGy is administered and noise with Hybrid-IR measures 15.9. To maintain approximately the same noise level with AI DLR, noise target would need to be set to SD = 35 and a CDTI of 1.5 mGy would be administered. At this dose level, the noise in the AI-DLR image was measured to be 16.1. Similarly, with noise target set at SD = 35, a CTDI of 1.5 mGy is administered and noise with Hybrid-IR measures 18.0. To maintain approximately the same noise level with AI DLR, noise target would need to be set to SD = 55 and a CDTI of 0.66 mGy would be administered. At this dose level, the noise in the AI-DLR image was measured to be 18.1. These results indicate a dose reduction of between 37% and 56% could be implemented without any noise penalty in the AI DLR images.

**Figure 5.**
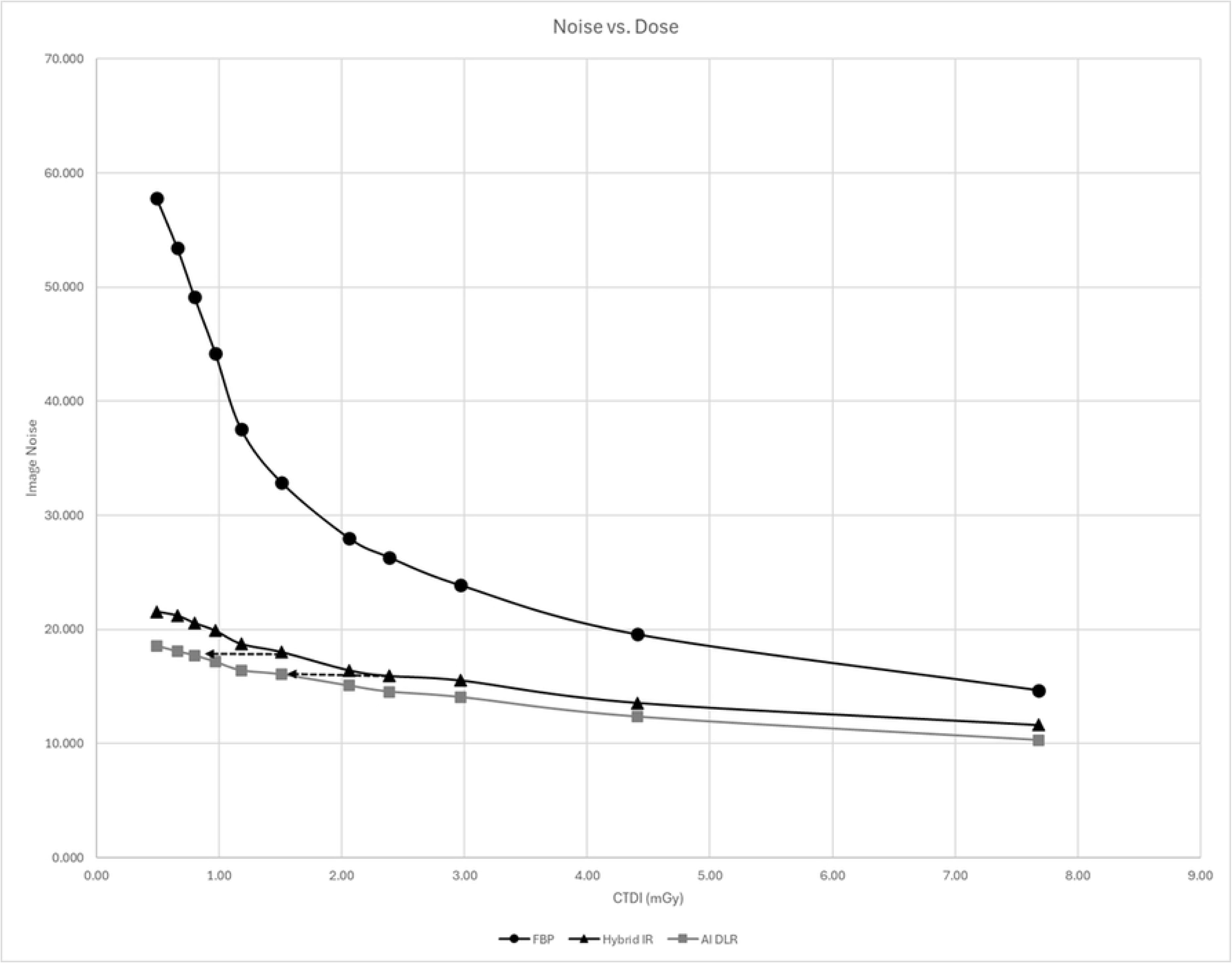
Impact of reconstruction algorithm on image noise. An equivalent noise level may be realized with AI DLR, but with greater than 36% lower dose compared to Hybrid-IR.

### QUALITATIVE

Qualitative image quality differences were consistent across all patients (Figure 6). The AI DLR image quality was mildly improved (+1 total difference) at soft tissue contrast and moderately improved (+2 total difference) at lung contrast compared with the Hybrid-IR. The Hybrid-IR image quality was moderately improved (+2 total difference) at soft tissue contrast and moderately (+2 total difference) at lung contrast compared with the FBP. The AI DLR was significantly improved (+3 total difference) at soft tissue contrast and even more significantly improved (+4 total difference) at lung contrast compared with the FBP.

**Figure 6.**
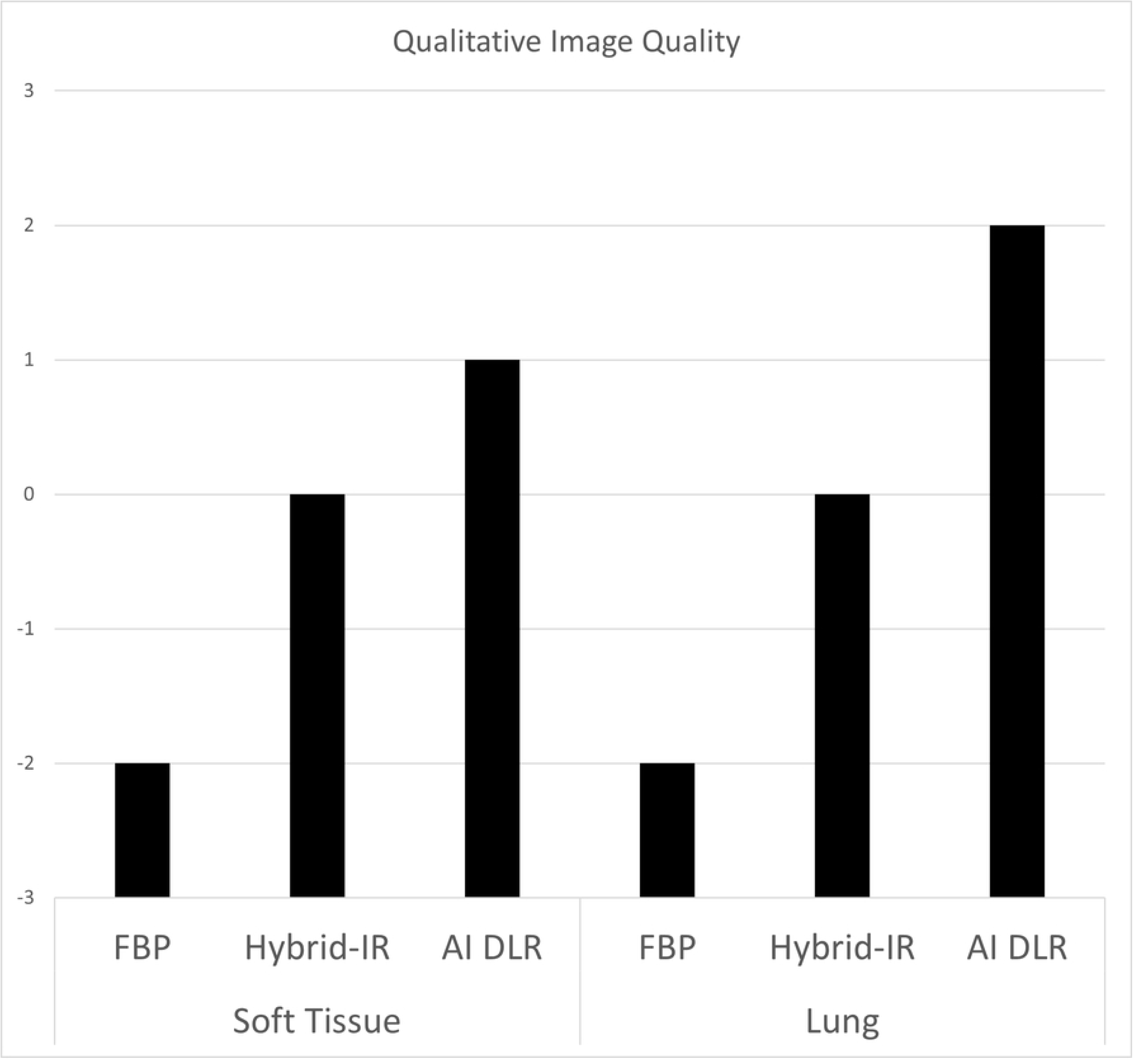
Relative image quality scores from qualitative image quality assessment.

## DISCUSSION

The results of this study quantitatively and qualitatively validated the use of AI Deep Learning Reconstruction for Coronary Artery Calcification Scoring and established that CACS can be consistently and robustly performed with AI DLR. Such validation results has yet to be reported for a 4 cm detector CT.

The CACS results with DLR show no statistically significant difference in Agaston values from those produced with those of a previously validated Hybrid-IR technique. However, they are generally marginally higher with AI DLR (mean 93.50) than with Hybrid-IR (mean 91.50). Possible reason for the differences between AI DLR and FBP is the lower noise in AI DLR removes a positive bias in CACS measurements caused by noise, while the improved spatial resolution improves the density measurements of small segments of calcium. These results are consistent with those previously reported [20].

There are two phenomena at play when AI DLR is used in the imaging of calcifications, and especially areas of small calcium deposits. AI DLR images are sharper, so there is less calcium blooming and smaller calcium deposits are detected more robustly. Secondarily, image noise is lower and as a result systematic biases in measurements are reduced. We believe the higher noise with FBP leads to more false positive and hence fewer cases classified as CACS = 0 with FBP compared to Hybrid-IR and AI DLR. Additionally, we believe the increased spatial resolution of AI-DLR is responsible for more tiny calcifications being detected, explaining the slightly fewer CAC=0 cases compared to Hybrid-IR.

This is supported by qualitative image analysis where individual calcifications may appear sharper and denser on AI DLR and FBP than on Hybrid-IR (Figure 7). It should also be noted that the FBP images have significantly worse clinical diagnostic quality than the AI DLR images and would not be advised for routine clinical use at either soft tissue or lung contrast settings. This is noted in the kappa agreement statistic for minimal calcium classification, which indicates only fair agreement (κ = 0.27 ± 0.14) between the hybrid IR and FBP reconstructions.

**Figure 7.**
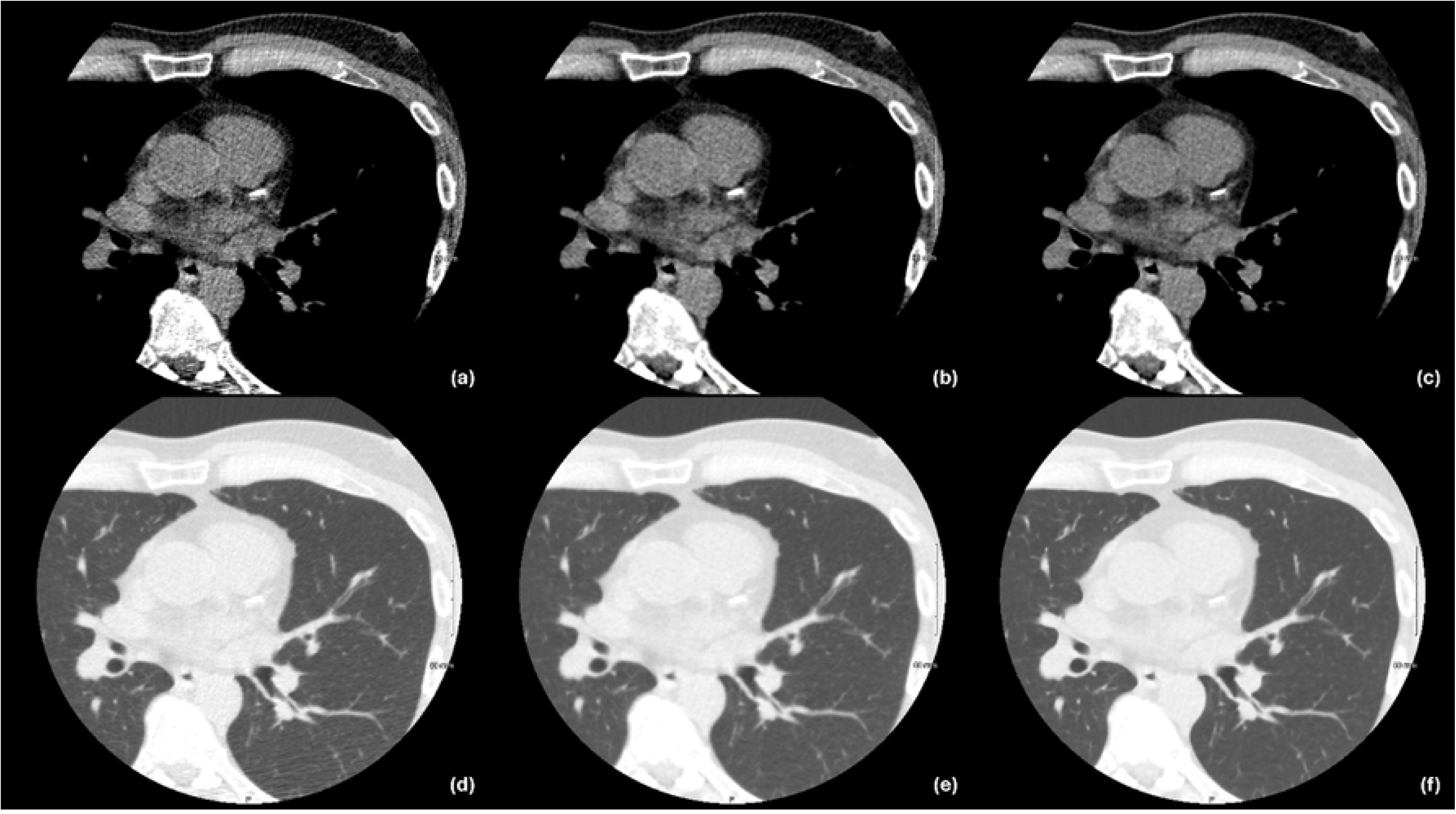
Example patient images displayed in Soft Tissue (WW = 350, WL = 40) and Lung (WW = 1600, WL = −650) windows. (a) Filter Back Projection (b) Hybrid-IR (c) AI DLR in Soft Tissue window; (d) Filter Back Projection (e) Hybrid-IR (f) AI DLR in Lung window. Note the calcifications are sharpest with AI DLR. Noise and artifacts are highest with FBP.

In general, there is substantial or near perfect agreement between all the AI DLR reconstructions and the Hybrid-IR reconstructions which is the current standard of care. Patients with minimal calcium classification, did show substantial agreement with κ = 0.64 ± 0.14. Lower measurements of calcium are more susceptible to noise in the measurements. It should be noted that interscan variability exists in CACS measurements. A number of factors including the choice of scanner can affect interscan variability, and the level of agreement in our study is less than variability that could be expected in general [24].

As reported in the results, patient dose was exponentially related to BMI Figure 3. This is consistent with previously studies investigating automated exposure control behavior [25].

AI DLR allows further dose reduction beyond the 1 mSv threshold set in this study for non-obese patients. Utilizing automated exposure control noise targets based on an AI DLR model rather than on Hybrid-IR model will allow for dose reductions of approximately 36% with improved image quality [26]. Further dose reduction can then be accomplished by utilizing a higher standard deviation of the noise (above 45) as the AI DLR image quality was improved compared with the Hybrid-IR image quality [20].

In conclusion, further dose reduction can be accomplished while maintaining image quality at or better than at current Hybrid-IR levels. We will target a dose reduction of at least a 37% for all patients. Accepting a linear, no threshold model of radiation risk [9, 27, 28] this we expect a similar reduction of radiation risk for all patients.

## TAKE HOME POINTS

1. AI DLR has been quantitatively and qualitatively validated for use in CACS on 4 cm coverage CT scanner.
2. AI DLR produces qualitatively better image quality than HYBRID-IR at the same dose level, while producing good agreement in categorization with Agatston scores.
3. AI DLR will allow for at least a 37% further dose reduction on a 4 cm coverage CT scanner.

## Data Availability

All data is stored on the personal computers of the authors. All data is scrubbed of all patient identification.

## BIBLIOGRAHY

1. Malguria N, Zimmerman S, Fishman EK. Coronary Artery Calcium Scoring: Current Status and Review of Literature. J Comput Assist Tomogr. 2018 Nov/Dec;42(6):887-897. doi: 10.1097/RCT.0000000000000825. PMID: 30422915.

2. Chang SM, Nabi F, Xu J, Pratt CM, Mahmarian AC, Frias ME, Mahmarian JJ. Value of CACS compared with ETT and myocardial perfusion imaging for predicting long-term cardiac outcome in asymptomatic and symptomatic patients at low risk for coronary disease: clinical implications in a multimodality imaging world. JACC Cardiovasc Imaging. 2015 Feb;8(2):134–44. doi: 10.1016/j.jcmg.2014.11.008. PMID: 25677886.

3. Budoff, M, Shaw, L, Liu, S. et al. Long-Term Prognosis Associated With Coronary Calcification: Observations From a Registry of 25,253 Patients. JACC. 2007 May, 49 (18) 1860–1870. 10.1016/j.jacc.2006.10.079

4. Sarwar A, Shaw LJ, Shapiro MD, Blankstein R, Hoffmann U, Cury RC, Abbara S, Brady TJ, Budoff MJ, Blumenthal RS, Nasir K. Diagnostic and prognostic value of absence of coronary artery calcification. JACC Cardiovasc Imaging. 2009 Jun;2(6):675–88. doi: 10.1016/j.jcmg.2008.12.031. Erratum in: JACC Cardiovasc Imaging. 2010 Oct;3(10):1089. Hoffman, Udo. PMID: 19520336.

5. McCollough CH, Ulzheimer S, Halliburton SS, Shanneik K, White RD, Kalender WA. Coronary artery calcium: a multi-institutional, multi-manufacturer international standard for quantification at cardiac CT. Radiology. 2007 May;243(2):527–38. doi: 10.1148/radiol.2432050808. PMID: 17456875.

6. Shreya D, Zamora DI, Patel GS, Grossmann I, Rodriguez K, Soni M, Joshi PK, Patel SC, Sange I. Coronary Artery Calcium Score - A Reliable Indicator of Coronary Artery Disease? Cureus. 2021 Dec 3;13(12):e20149. doi: 10.7759/cureus.20149. PMID: 35003981; PMCID: PMC8723785.

7. Marwan M, Mettin C, Pflederer T, Seltmann M, Schuhbäck A, Muschiol G, Ropers D, Daniel WG, Achenbach S. Very low-dose coronary artery calcium scanning with high-pitch spiral acquisition mode: comparison between 120-kV and 100-kV tube voltage protocols. J Cardiovasc Comput Tomogr. 2013 Jan-Feb;7(1):32-8. doi: 10.1016/j.jcct.2012.11.004. Epub 2012 Dec 1. PMID: 23333186.

8. Cao CF, Ma KL, Shan H, Liu TF, Zhao SQ, Wan Y, Jun-Zhang, Wang HQ. CT Scans and Cancer Risks: A Systematic Review and Dose-response Meta-analysis. BMC Cancer. 2022 Nov 30;22(1):1238. doi: 10.1186/s12885-022-10310-2. PMID: 36451138; PMCID: PMC9710150.

9. Little, M.P., Wakeford, R., Tawn, E.J., Bouffler, S.D., & De Gonzalez, A.B. (2009). Risks Associated with Low Doses and Low Dose Rates of Ionizing Radiation: Why Linearity May Be (Almost) the Best We Can Do. Radiology, 251(1), 6–12. 10.1148/radio.2511081686

10. Einstein AJ, Henzlova MJ, Rajagopalan S. Estimating risk of cancer associated with radiation exposure from 64-slice computed tomography coronary angiography. JAMA. 2007 Jul 18;298(3):317–23. doi: 10.1001/jama.298.3.317. PMID: 17635892

11. BE. Earth, D.O. (2006). Health Risks from Exposure to Low Levels of Ionizing Radiation: BEIR VII Phase 2. https://openlibrary.org/books/OL18209291M/Health_risks_from_exposure_to_low_levels_of_ionizing_radiation_BEIR_VII_Phase_2.

12. Choi AD, Leifer ES, Yu JH, Datta T, Bronson KC, Rollison SF, Schuzer JL, Steveson C, Shanbhag SM, Chen MY. Reduced radiation dose with model based iterative reconstruction coronary artery calcium scoring. Eur J Radiol. 2019 Feb;111:1–5. doi: 10.1016/j.ejrad.2018.12.010. Epub 2018 Dec 8. PMID: 30691659.

13. Takahashi M, Kimura F, Umezawa T, Watanabe Y, Ogawa H. Comparison of adaptive statistical iterative and filtered back projection reconstruction techniques in quantifying coronary calcium. J Cardiovasc Comput Tomogr. 2016 Jan-Feb;10(1):61-8. doi: 10.1016/j.jcct.2015.07.012. Epub 2015 Jul 29. PMID: 26276567.

14. Choi AD, Leifer ES, Yu J, Shanbhag SM, Bronson K, Arai AE, Chen MY. Prospective evaluation of the influence of iterative reconstruction on the reproducibility of coronary calcium quantification in reduced radiation dose 320 detector row CT. J Cardiovasc Comput Tomogr. 2016 Sep-Oct;10(5):359-63. doi: 10.1016/j.jcct.2016.07.016. Epub 2016 Jul 27. PMID: 27591767; PMCID: PMC7458582.

15. Tatsugami F, Higaki T, Fukumoto W, Kaichi Y, Fujioka C, Kiguchi M, Yamamoto H, Kihara Y, Awai K. Radiation dose reduction for coronary artery calcium scoring at 320-detector CT with adaptive iterative dose reduction 3D. Int J Cardiovasc Imaging. 2015 Jun;31(5):1045–52. doi: 10.1007/s10554-015-0637-7. Epub 2015 Mar 10. PMID: 25754302.

16. Chen MY, Steigner ML, Leung SW, Kumamaru KK, Schultz K, Mather RT, Arai AE, Rybicki FJ. Simulated 50 % radiation dose reduction in coronary CT angiography using adaptive iterative dose reduction in three-dimensions (AIDR3D). Int J Cardiovasc Imaging. 2013 Jun;29(5):1167–75. doi: 10.1007/s10554-013-0190-1. Epub 2013 Feb 13. PMID: 23404384; PMCID: PMC3701132.

17. Yin WH, Lu B, Li N, Han L, Hou ZH, Wu RZ, Wu YJ, Niu HX, Jiang SL, Krazinski AW, Ebersberger U, Meinel FG, Schoepf UJ. Iterative reconstruction to preserve image quality and diagnostic accuracy at reduced radiation dose in coronary CT angiography: an intraindividual comparison. JACC Cardiovasc Imaging. 2013 Dec;6(12):1239–49. doi: 10.1016/j.jcmg.2013.08.008. Epub 2013 Oct 23. PMID: 24269265.

18. Tatsugami F, Higaki T, Nakamura Y, Yu Z, Zhou J, Lu Y, Fujioka C, Kitagawa T, Kihara Y, Iida M, Awai K. Deep learning-based image restoration algorithm for coronary CT angiography. Eur Radiol. 2019 Oct;29(10):5322–5329. doi: 10.1007/s00330-019-06183-y. Epub 2019 Apr 8. PMID: 30963270

19. Chen MY, Steigner ML, Leung SW, Kumamaru KK, Schultz K, Mather RT, Arai AE, Rybicki FJ. Simulated 50 % radiation dose reduction in coronary CT angiography using adaptive iterative dose reduction in three-dimensions (AIDR3D). Int J Cardiovasc Imaging. 2013 Jun;29(5):1167–75. doi: 10.1007/s10554-013-0190-1. Epub 2013 Feb 13. PMID: 23404384; PMCID: PMC3701132.

20. Otgonbaatar C, Jeon PH, Ryu JK, Shim H, Jeon SH, Ko SM, Kim H. Coronary artery calcium quantification: comparison between filtered-back projection, hybrid iterative reconstruction, and deep learning reconstruction techniques. Acta Radiol. 2023 Aug;64(8):2393–2400. doi: 10.1177/02841851231174463. Epub 2023 May 21. PMID: 37211615.

21. Higaki T, Nakamura Y, Zhou J, Yu Z, Nemoto T, Tatsugami F, Awai K. Deep Learning Reconstruction at CT: Phantom Study of the Image Characteristics. Acad Radiol. 2020 Jan;27(1):82–87. doi: 10.1016/j.acra.2019.09.008. PMID: 31818389.

22. American Association of Physicists in Medicine Report # 96; The Measurement, Reporting, and Management of Radiation dose in CT; Report of AAPM task Group 23 of the Diagnostic Imaging Council CT Committee; January, 2008

23. Mitani AA, Freer PE, Nelson KP. Summary measures of agreement and association between many raters’ ordinal classifications. Ann Epidemiol. 2017 Oct;27(10):677–685.e4. doi: 10.1016/j.annepidem.2017.09.001. Epub 2017 Sep 22. PMID: 29029991; PMCID: PMC5687310.

24. Kaitlin B. Baron KB, Choi AD, Chen MY. Low Radiation Dose Calcium Scoring: Evidence and Techniques. Curr Cardiovasc Imaging Rep. Epub 2016 March; (2016) 9:12; doi: 10.1007/s12410-016-9373-1

25. Inoue Y, Itoh H, Nagahara K, Hata H, Mitsui K. Relationships of Radiation Dose Indices with Body Size Indices in Adult Body Computed Tomography. Tomography. 2023 Jul 14;9(4):1381–1392. doi: 10.3390/tomography9040110. PMID: 37489478; PMCID: PMC10366833.

26. Hoe, J., & Tan, C. (2022, September) Advanced intelligent Clear-IQ Engine in Clinical Practice, White Paper. Canon Medical Systems. https://global.medical.canon/publication/ct/VS3_04_MOIGE0040EAB_SYMPOSIUM.

27. Little MP, Wakeford R, Tawn EJ, Bouffler SD, Berrington de Gonzalez A. Risks associated with low doses and low dose rates of ionizing radiation: why linearity may be (almost) the best we can do. Radiology. 2009 Apr;251(1):6–12. doi: 10.1148/radiol.2511081686. PMID: 19332841; PMCID: PMC2663578.

28. Mettler FA, Upton AC, Chapter 4 - Cancer Induction and Dose-Response Models, Editor(s): Fred A. Mettler, Arthur C. Upton, Medical Effects of Ionizing Radiation (Third Edition), W.B. Saunders, 2008, Pages 71–115.

